# AI-Driven Fluorescence Peak Analysis for Chromosomal Aneuploidy Detection: A Python-Based Machine Learning Approach for Enhanced Accuracy and Efficiency

**DOI:** 10.1101/2025.03.22.25324455

**Authors:** Krishna H. Goyani, Daisy Patel, Isha Sharma, Shalin Vaniawala, Pratap N. Mukhopadhyaya

**Affiliations:** Wobble Base Bioresearch Private limited, Pune; Parul University, Waghodia, Vadodara, Gujarat; New Civil Hospital, Jay Prakash Narayan Marg, Surat

**Keywords:** Chromosomal aneuploidy, trisomy detection, segmental duplication, QF-PCR, machine learning

## Abstract

Chromosomal aneuploidy, a condition characterized by an abnormal number of chromosomes, is a major genetic disorder affecting human reproduction, leading to infertility, pregnancy loss, and developmental disabilities. Trisomies of chromosomes 13, 18, and 21 result in Patau, Edwards, and Down syndromes, respectively. While conventional methods like karyotyping and QF-PCR facilitate aneuploidy detection, they are often time-consuming and limited by genetic polymorphism variability. This study introduces an advanced AI-driven approach integrating segmental duplication-based fluorescence probe analysis with machine learning for efficient and accurate aneuploidy detection. Amniotic fluid samples were collected from pregnant mothers, and DNA was extracted for QF-PCR amplification of segmental duplications on target chromosomes. Fluorescence intensity data were analyzed using a Python-based computational pipeline employing an XGBoost classifier trained on 80% of the dataset and tested on the remaining 20%. The model demonstrated high accuracy in detecting trisomies 13, 18, and 21, with results validated against conventional karyotyping as the gold standard. Furthermore, the AI-based approach successfully predicted fetal gender by computing fluorescence intensity ratios of X and Y chromosomes relative to reference chromosomes. The automated method significantly reduced analysis time from 45 minutes (manual interpretation) to 1.7 seconds while minimizing human errors. The integration of two reference chromosomes for fluorescence normalization improved diagnostic precision, reducing false positives and negatives. This study highlights the potential of AI-enhanced QF-PCR analysis for rapid and reliable prenatal aneuploidy screening, paving the way for its implementation in clinical diagnostics to enhance reproductive healthcare outcomes.

## Introduction

Chromosomal aneuploidy, characterized by an abnormal number of chromosomes, is a significant genetic disorder in humans with far-reaching implications. It is a common cause of infertility, occurring in approximately 15% of couples trying to conceive (Harton & Tempest, 2011). Aneuploidy is also the leading cause of pregnancy loss and developmental disabilities, with over 25% of all miscarriages being monosomic or trisomic (Hassold et al., 1996). Furthermore, it is present in an estimated 10-30% of all fertilized eggs, making it a major factor in human reproduction and development (Hassold et al., 1996). Interestingly, the consequences of aneuploidy are not always straightforward. While it is generally detrimental, some studies have shown that aneuploid embryos can serve as a source for both normal euploid and aneuploid human embryonic stem cell (hESC) lines (Biancotti et al., 2010). These cell lines can be invaluable tools for studying developmental aspects of chromosomal abnormalities in humans. Additionally, in the context of cancer, aneuploidy has been found to have a complex relationship with tumorigenesis. Despite its frequency in human tumors, aneuploidy is not always a driver of cancer development and can even exert tumor-suppressive effects in some cases (Vasudevan et al., 2021).

Aneuploidy in chromosomes 13, 18, and 21 results in Patau syndrome, Edwards syndrome, and Down syndrome, respectively. These are the only full autosomal trisomies compatible with postnatal survival (Altug-Teber et al., 2008). The mechanisms underlying the disruption of normal development and specific phenotypes in these syndromes are not fully understood, but research suggests a combination of gene dosage effects and genome-wide transcriptional dysregulation (Altug-Teber et al., 2008; Hwang et al., 2021). Interestingly, the transcriptional changes vary among the different trisomies. In trisomy 21, a subset of chromosome 21 genes, including DSCR1 involved in fetal heart development, shows consistent up-regulation, while trisomy 18 exhibits more extensive downstream transcriptional changes (Altug-Teber et al., 2008). Additionally, aneuploidy-associated phenotypes, such as lower viability and increased dependency on serine-driven lipid synthesis, are present in trisomy 21 cells, independent of the identity of the triplicated genes (Hwang et al., 2021).

QF-PCR offers a rapid, precise, and automated method capable of handling 96 samples in less than 48 hours. Nonetheless, its drawback lies in the variability of genetic polymorphisms among different populations, which limits its universal applicability (Atef et al., 2011; Slater et al., 2003; Dudarewicz et al., 2005). On the other hand, MLPA is a validated technique for identifying changes in genomic copy numbers and is used in aneuploidy analysis. However, it involves an overnight hybridization step, which makes the process time-consuming and complex to develop (Boormans et al., 2010; Willis et al., 2012).

Segmental duplications play a crucial role in detecting human chromosomal aneuploidy and other structural abnormalities. These duplications are regions of DNA that are repeated within the genome and can serve as markers for identifying chromosomal aberrations (Bailey et al., 2002).

In this study, the concept of segmental duplication was exploited to integrate it with a fragment analysis protocol running on a genetic analyzer, where relative dosage is computed after comparing the signal generated from the target chromosome with that of two reference chromosomes (Kong et al., 2014). Python-based code was utilized to analyze QF-PCR data for human aneuploidy. To automate data interpretation, a model was trained on 70% of the dataset and tested on the remaining 30%, ensuring robust validation of the analytical approach. This computational framework enhances the efficiency and accuracy of aneuploidy detection, minimizing manual errors and improving reproducibility.

## Material and methods

### Clinical Samples and DNA Extraction

Clinical samples comprised amniocentesis fluid collected from pregnant mothers. DNA was extracted using the QIAamp DNA Mini Kit (Qiagen, Hilden, Germany) at a NABL-accredited laboratory (ISO 15189:2022 for medical testing). All extracted DNA samples were stored at −20°C until further processing. The quality and quantity of the extracted DNA were assessed using a NanoDrop spectrophotometer (NanoDrop™ 2000, Thermo Fisher Scientific, USA), measuring absorbance at 260 nm and 280 nm to determine purity and concentration.

### PCR Amplification and Fragment Analysis

Approximately 50 ng of DNA extracted from amniocentesis fluid from each patient was used to set up a PCR reaction. The reaction was performed in a 25 μL volume containing 1X PCR buffer, 200 μM dNTPs, 1.5 mM MgCl□, 0.5 U Taq DNA polymerase (Thermo Fisher Scientific, USA), and 0.2 μM each of forward and reverse primers, where the forward primer was fluorescently labeled. PCR was carried out under the following conditions: initial denaturation at 95°C for 5 minutes, followed by 35 cycles of denaturation at 95°C for 30 seconds, annealing at an optimized temperature (specific to primers) for 30 seconds, and extension at 72°C for 30 seconds, with a final extension at 72°C for 7 minutes. The PCR products were analyzed using a 3500 Genetic Analyzer (Applied Biosystems, USA), and fragment analysis was performed using GeneMapper software v6.0 (Thermo Fisher Scientific, USA). The sequences of the PCR primers were as described by Kong et al., 2014. The primers enabled the simultaneous detection of aneuploidies. Two independent primer sets (targeting two segmental duplications per chromosome) were employed. Each primer pair included one unlabelled primer and one labeled with FAM (6-carboxyfluorescein). All PCR primers were synthesized and purified by Thermo Fisher Scientific, India.

### Computational Analysis and Machine Learning Implementation

Fluorescence intensity data obtained from experimental assays were processed using a Python-based computational pipeline. The dataset, stored in CSV format, contained fluorescence intensity values for target chromosomes (16, 18, 21, X, Y) and reference chromosomes, along with labels indicating chromosomal aneuploidy status. Preprocessing was performed using pandas and numpy, including data inspection, categorical label encoding via LabelEncoder, and normalization of fluorescence intensity values. The dataset was partitioned into training (80%) and testing (20%) subsets using train_test_split from scikit-learn to ensure unbiased model evaluation.

For automated classification, an XGBoost classifier was implemented, leveraging its gradient boosting framework for efficient learning. The model was initialized with a multi:softmax objective for multiclass classification, mlogloss as the evaluation metric, and five output classes corresponding to chromosomal categories. Training was conducted using fluorescence intensity features as predictors and encoded chromosome labels as targets. The fit method of XGBClassifier was applied to train the model on the prepared dataset. Model predictions were obtained using predict on the test subset, and performance evaluation was carried out using classification reports (classification_report) and ROC-AUC scores (roc_auc_score) with a one-vs-rest (OVR) approach.

Additionally, fluorescence intensity ratios of chromosomes X and Y relative to a reference chromosome were computed to predict fetal gender. These ratios were incorporated as model features, allowing the AI-driven classifier to distinguish between male and female samples. The entire workflow, including dataset handling, feature engineering, model training, and evaluation, was executed within a Python environment utilizing pandas, numpy, xgboost, and sklearn for efficient machine learning-based analysis. All relevant artifacts, including datasets in Google Sheets format and Python code as Colab notebooks, were compiled in a Cloud Storage folder for access and sharing.

### Gold Standard Confirmation

Karyotyping was performed as the gold standard for confirming trisomy in chromosomes 13, 18, and 21. Peripheral blood samples were cultured in RPMI-1640 medium supplemented with fetal bovine serum and phytohemagglutinin for 72 hours. Metaphase chromosomes were arrested using colchicine, followed by hypotonic treatment and fixation with methanol-acetic acid. Chromosomal spreads were stained with Giemsa and analyzed under a light microscope. Karyotypes were classified according to ISCN guidelines, and trisomy cases identified via QF-PCR were cross-validated against karyotyping results to ensure diagnostic accuracy.

### Ethics Statement

This study was approved by the Wobble Base Bioresearch Ethics Committee (Approval No. WBBPL/EC/Jan2005/002) and conducted in accordance with the ethical guidelines outlined in the Declaration of Helsinki. Written informed consent was obtained from all participants or their legal guardians prior to sample collection and analysis.

### Age Statement

The participants in this study ranged in age from 18 to 49 years, ensuring inclusion of reproductive-age women. Age data are presented in five-year range intervals to maintain confidentiality.

## Results

For each sample, data were collected from one or more of the five target chromosomes: 13, 18, 21, X, and Y. In samples where both the X and Y chromosomes were analyzed, the genetic sex was predicted. Each target chromosome was analyzed in parallel with a pair of reference chromosomes to ensure accurate comparison. The reference chromosomes used were as follows: Chromosome 21 with Chromosomes 11 and 6, Chromosome 18 with Chromosomes 10 and 1, Chromosome 13 with Chromosomes 11 and 9, Chromosome Y with Chromosome X, and Chromosome X with Chromosomes 3 and 18. The data capture value was determined based on the fluorescent peak obtained from the GeneMapper software.

The table provided in the supplement data section (<u>Table (Supplement data)</u>.) presents details of the total number of clinical samples processed and the distribution of samples tested for specific chromosomal aneuploidies, including trisomy 21 (Down syndrome), trisomy 18 (Edwards syndrome), and trisomy 13 (Patau syndrome). The numbers indicate samples tested for each trisomy individually or in combination with other trisomies. Data interpretation was conducted using a Python-based machine learning algorithm for automated analysis, with standard karyotyping serving as the gold standard for validation.

Table 1 presents the distribution of samples processed and tested for chromosomal aneuploidies. A total of 142 samples were analyzed, with 139 tested for trisomy 21 (Down syndrome), 95 for trisomy 18 (Edwards syndrome), and 93 for trisomy 13 (Patau syndrome), either individually or in combination with other trisomies. Additionally, 92 samples underwent comprehensive chromosomal analysis. Data interpretation was conducted using a Python-based artificial intelligence (AI) algorithm, with standard karyotyping serving as the gold standard for validation.

**Table 1.** Summary of Sample Testing for Chromosomal Aneuploidy

| **Description** | **Count** |
| --- | --- |
| Total number of samples processed | 142 |
| Number of samples tested for trisomy 21 (alone or along with trisomy 13 and/or 18) | 139 |
| Number of samples tested for trisomy 18 (alone or along with trisomy 13 and/or 21) | 95 |
| Number of samples tested for trisomy 13 (alone or along with trisomy 18 and/or 21) | 93 |
| Number of samples tested for all the chromosomes | 92 |

In Table 2, a comparative analysis of chromosomal aneuploidy detection using an AI-based interpretation method versus conventional karyotyping is presented. The Count (AI) column represents the number of samples identified as positive for trisomy 21, trisomy 18, and trisomy 13 by the Python-based AI process, while the Count (Gold standard) column indicates the corresponding results obtained through standard karyotyping. Discrepancies, if any, highlight potential variations in detection sensitivity between the bench data driven AI-based approach and the established cytogenetic method.

**Table 2.** Comparison of AI-Based Interpretation and Gold Standard Karyotyping for Trisomy Detection

| Description | Count (AI) | Count (Gold standard) |
| --- | --- | --- |
| Number of samples found positive for trisomy 21 | 12 | 13 |
| Number of samples found positive for trisomy 18 | 3 | 3 |
| Number of samples found positive for trisomy 13 | 0 | 0 |

*AI: Artificial Intelligence*

Table 3 summarizes the determination of gender chromosomes in 92 processed samples using an AI-based interpretation method and conventional karyotyping. The Count (AI) column represents the classification results from the Python-based AI process, while the Count (Gold standard) column indicates the corresponding results obtained through standard karyotyping. Both methods yielded identical classifications, with 43 samples identified as male and 49 as female, demonstrating concordance between AI-based analysis and cytogenetic evaluation.

**Table 3.** AI-Based Interpretation and Gold Standard Karyotyping for Gender Chromosome Determination.

| **Description** | **Count**<br>**(AI)** | **Count**<br>**(Gold**<br>**standard)** |
| --- | --- | --- |
| Total number of samples processed for determining the gender chromosomes | 92 | 92 |
| Number of male samples | 43 | 43 |
| Number of female samples | 49 | 49 |

The machine learning approach completed the analysis in 1.7 seconds, compared to the 45 minutes required for manual analysis. Notably, the AI-based method successfully detected and accurately highlighted an error originating from a bench-level variable, which might have been overlooked in manual interpretation.

As mentioned above, this study utilized an AI-driven approach to analyze trisomy-related fluorescence PCR data. Fluorescence intensity ratios were computed for chromosomes X and Y relative to a reference chromosome to predict fetal gender. These ratios were integrated as model features, enabling the classifier to distinguish between male and female samples. The entire workflow—spanning dataset preprocessing, feature extraction, model training, and evaluation—was conducted in a Python environment using pandas, numpy, xgboost, and sklearn, ensuring efficient machine learning-based analysis.

## Discussion

Detecting trisomy in pregnant mothers is crucial for several reasons: Trisomy 21 (Down syndrome) is the most common reason women opt for prenatal diagnosis (Lo et al., 2007). Early detection allows parents to make informed decisions about pregnancy management and prepare for potential medical needs. Conventional invasive methods like amniocentesis carry risks, driving the development of noninvasive techniques (Chen et al., 2011; Lo et al., 2007). Interestingly, while trisomy screening has advanced significantly, some studies show that a small percentage of parents choose to continue pregnancies even after trisomy diagnosis. For instance, 12% of couples continued pregnancies after confirming trisomy 13 or 18 diagnoses (Parker et al., 2003). In summary, trisomy detection enables early intervention, informed decision-making, and preparation for potential medical needs. The development of noninvasive prenatal testing (NIPT) has made screening more accessible and safer, allowing for earlier and more widespread detection of chromosomal abnormalities during pregnancy (Benn, 2014; Zheng et al., 2020). This study attempted an important technological advancement relevant to a prominent health condition in pregnant females.

Segmental duplications can be used as a tool for detecting trisomy, a chromosomal abnormality where an extra copy of a chromosome is present. Array comparative genomic hybridization (CGH) is a technique that can detect these duplications and, by extension, trisomy (Chen et al., 2005). This method compares the DNA of a test sample against a reference genome, allowing for the identification of copy number variations, including large-scale duplications characteristic of trisomy. Interestingly, segmental duplications themselves can lead to chromosomal rearrangements and copy number variations. For instance, in Charcot-Marie-Tooth disease type 1A, a segmental duplication on chromosome 17p is linked to the condition (Kaku et al., 1993). This highlights the dual nature of segmental duplications – they can be both a cause of genomic variation and a tool for detecting it. While segmental duplications can be used to detect trisomy through techniques like array CGH, they also play a complex role in genome evolution and disease. The study of these duplications provides insights into chromosomal abnormalities and genomic variation (Cheng et al., 2005; Johnson et al., 2006). In this study, segmental duplication was strategically used to detect chromosome dosage in target chromosomes (Chromosomes 13, 18, and 21) using fluorescence probes, endpoint PCR, and fragment analysis on a genetic analyzer.

In this study, the use of two reference chromosomes for determining the dosage of target chromosomes (Chromosomes 13, 18, and 21) in segmental duplication-based fluorescence probe analysis and fragment analysis using a genetic analyzer provided several advantages. Incorporating two references improved accuracy by offering a stable baseline for comparison, minimizing the impact of experimental variability. This approach also reduced the risk of false positives and negatives, as reliance on a single reference could introduce bias due to amplification inconsistencies. By normalizing fluorescence intensity variations, the use of dual reference chromosomes ensured greater consistency in dosage assessment. Additionally, this strategy enhanced the statistical confidence in detecting trisomy conditions, particularly in borderline cases. The inclusion of two references also mitigated potential errors arising from undetected structural variations or aneuploidy in a single reference chromosome. Furthermore, in fluorescence-based fragment analysis, dual references provided refined calibration, improving the distinction between normal and trisomic samples.

Python-based environments are widely used for biological data analysis, from dataset preprocessing and feature engineering to model training and evaluation. This approach offers several advantages in handling complex biological datasets. Data preprocessing and feature engineering are crucial steps in biological data analysis. For instance, in microbiome data analysis, compositional transformations and filtering methods are often employed, although their impact on predictive performance can vary (Papoutsoglou et al., 2023). In the context of high-dimensional biological datasets, feature selection techniques like the Statistically Equivalent Signatures algorithm have proven effective in reducing classification errors (Papoutsoglou et al., 2023). For biological feature selection, metaheuristic algorithms such as the general learning equilibrium optimizer (GLEO) have shown excellent performance in identifying informative features among a large number of attributes (Too & Mirjalili, 2020). Interestingly, some studies have found that certain preprocessing techniques may not always improve model performance. For example, in microbiome data analysis, the use of compositional transformations and filtering methods did not consistently enhance predictive performance (Papoutsoglou et al., 2023). This highlights the importance of carefully evaluating preprocessing steps in the context of specific biological datasets and research questions. Python-based environments hence offer powerful tools for biological data analysis, from preprocessing to model evaluation. The choice of preprocessing techniques and feature selection methods should be tailored to the specific characteristics of the biological dataset and the research objectives. Techniques like multivariate feature selection and metaheuristic algorithms have shown promise in improving model performance and biological insights (Papoutsoglou et al., 2023; Too & Mirjalili, 2020). However, it’s crucial to critically evaluate the impact of preprocessing steps on model performance and biological interpretability.

The samples originated from a standard genetic testing laboratory, where the choice of the trisomy detection protocol was driven by the specific research and development program underway, as per organizational requirements. This resulted in heterogeneity in the testing approach, with some samples being analyzed for all three target chromosomes (21, 18, and 13), while others were tested for only one or two of these trisomies.

Trisomy 21 is the most common chromosomal disorder among live births. It occurs in approximately 1 in 700 to 1 in 1,000 live births worldwide (Sánchez-Pavón et al., 2022). This genetic condition results from the presence of an extra copy of chromosome 21, leading to various developmental abnormalities and intellectual disability (Cooper et al., 2012; Hibaoui et al., 2013). Interestingly, while maternal age is the primary risk factor for trisomy 21, recent studies have shown that paternal age and epigenetic factors also play a role in its occurrence (Sánchez-Pavón et al., 2022). Additionally, research has revealed that in prenatal diagnoses, the paternal origin of trisomy 21 is more frequent (10.8%) than previously thought based on studies of liveborn infants (6.7%) (Muller et al., 2000). This suggests a potential impact of fetal death on the observed frequencies of parental origin in trisomy 21 cases. In conclusion, trisomy 21 remains the most prevalent chromosomal abnormality, with its abundance attributed to various factors including parental age and epigenetic influences. The discrepancy in paternal origin frequencies between prenatal and postnatal studies highlights the complexity of this condition and the need for further research to fully understand its etiology and prevalence patterns. Our study corroborates this observation, as trisomy 21 was found to be the most prevalent among the three target chromosomes investigated in this study, namely chromosomes 13, 18, and 21.

In our study, the segmental duplication-based approach for fetal gender determination demonstrated perfect accuracy. The dosage obtained from the X and Y chromosomes in male fetuses, as well as the dosage derived from a pair of X chromosomes in female fetuses, when analyzed against reference chromosomes, reliably identified fetal gender.

This study leveraged an AI-driven approach to analyze fluorescence PCR data related to trisomy detection, incorporating fluorescence intensity ratios of chromosomes X and Y relative to a reference chromosome for fetal gender prediction. By integrating these ratios as model features, the classifier effectively distinguished between male and female samples. The entire computational pipeline—from dataset preprocessing and feature extraction to model training and evaluation—was implemented in a Python-based environment using *pandas, numpy, xgboost*, and *sklearn*, ensuring a streamlined and efficient analysis. Notably, the AI-based method achieved an approximately 1,588-fold improvement in processing speed compared to manual analysis, underscoring its superiority in handling large-scale genomic datasets with enhanced accuracy and minimal human intervention.

## Conclusion

This study demonstrates the efficacy of an AI-driven approach for trisomy detection and fetal gender prediction using fluorescence PCR data. By integrating machine learning techniques with fluorescence intensity ratio analysis and dual-reference chromosome normalization, the method achieved superior accuracy and significantly reduced processing time compared to manual analysis. The use of Python-based computational frameworks enabled efficient data preprocessing, feature selection, and model evaluation, highlighting the power of AI in genomic diagnostics. These findings emphasize the potential of AI-assisted noninvasive prenatal testing (NIPT) to improve trisomy screening, enhance diagnostic confidence, and expand accessibility to early genetic risk assessment in prenatal care. Future advancements in AI-driven genomic analysis may further refine detection accuracy and broaden applications in precision medicine.

## Supporting information

https://docs.google.com/document/d/1drvycX4dD-qW9Hb4SRQ9z6WlK2AEGfKy/edit?usp=sharing&ouid=115148606976539336609&rtpof=true&sd=true

## Acknowledgement

The authors sincerely acknowledge SN Gene Lab Private Limited, Gujarat, India, for granting access to their clinical sample repository and genetic analyzer for fragment analysis, which was essential in generating the dataset for this study. Furthermore, the authors express their gratitude to Indraneel Mukhopadhaya (Postgraduate in Applied Statistics and Informatics, Indian Institute of Technology, Bombay, India) for his insightful suggestions on the Python-based analytical aspects of this research.

