## Supplementary material for "AI-Driven Fluorescence Peak Analysis for Chromosomal Aneuploidy Detection: A Python-Based Machine Learning Approach for Enhanced Accuracy and Efficiency": https://docs.google.com/document/d/1drvycX4dD-qW9Hb4SRQ9z6WlK2AEGfKy/edit?usp=sharing&ouid=115148606976539336609&rtpof=true&sd=true

**[Table (Supplement data).](https://docs.google.com/document/d/1drvycX4dD-qW9Hb4SRQ9z6WlK2AEGfKy/edit?usp=sharing&ouid=115148606976539336609&rtpof=true&sd=true)** Results of quantitative fluorescence PCR (QF-PCR) analysis for detecting chromosomal aneuploidies, including trisomy 21 (Down syndrome), trisomy 18 (Edwards syndrome), and trisomy 13 (Patau syndrome).

| **Serial No.** | **Sample ID** | **Ref. Chr Name** | **Peak Height** | **Target Chr Name** | **Peak Height** | **AI Based Interpretation** | **Gold Standard (Karyotyping)** |
| --- | --- | --- | --- | --- | --- | --- | --- |
| 1 | 34587 | 11 | 1567 | 21 | 1568 | Normal | Normal |
|  |  | 6 | 1541 | 21 | 1519 |  |  |
|  |  | 10 | 1717 | 18 | 1521 | Normal | Normal |
|  |  | 1 | 1832 | 18 | 1904 |  |  |
|  |  | 11 | 1191 | 13 | 1097 | Normal | Normal |
|  |  | 9 | 1960 | 13 | 1628 |  |  |
|  |  | X | 997 | Y | 823 | Male | Male |
|  |  | X | 960 | Y | 1011 |  |  |
|  |  | 3 | 1410 | X | 669 |  |  |
|  |  | 18 | 1725 | X | 845 |  |  |
| 2 | 34800 | 11 | 2118 | 21 | 2183 | Normal | Normal |
|  |  | 6 | 1920 | 21 | 1959 |  |  |
|  |  | 10 | 2462 | 18 | 2091 | Normal | Normal |
|  |  | 1 | 2561 | 18 | 2516 |  |  |
|  |  | 11 | 1855 | 13 | 1912 | Normal | Normal |
|  |  | 9 | 2588 | 13 | 2106 |  |  |
|  |  | X | 2138 | Y |  | Female | Female |
|  |  | X | 2334 | Y |  |  |  |
|  |  | 3 | 1942 | X | 1820 |  |  |
|  |  | 18 | 2590 | X | 2621 |  |  |
| 3 | 34810 | 11 | 552 | 21 | 474 | Normal | Normal |
|  |  | 6 | 447 | 21 | 402 |  |  |
|  |  | 10 | 601 | 18 | 503 | Normal | Normal |
|  |  | 1 | 370 | 18 | 394 |  |  |
|  |  | 11 | 251 | 13 | 214 | Normal | Normal |
|  |  | 9 | 560 | 13 | 508 |  |  |
|  |  | X | 539 | Y |  | Female | Female |
|  |  | X | 312 | Y |  |  |  |
|  |  | 3 | 303 | X | 277 |  |  |
|  |  | 18 | 328 | X | 303 |  |  |
| 4 | 34729 | 11 | 5385 | 21 | 5080 | Normal | Normal |
|  |  | 6 | 4874 | 21 | 4911 |  |  |
| 5 | 34730 | 11 | 9371 | 21 | 9197 | Normal | Normal |
|  |  | 6 | 8636 | 21 | 8034 |  |  |
| 6 | 34926 | 11 | 812 | 21 | 806 | Normal | Normal |
|  |  | 6 | 921 | 21 | 889 |  |  |
|  |  | 10 | 986 | 18 | 992 | Normal | Normal |
|  |  | 1 | 603 | 18 | 628 |  |  |
|  |  | 11 | 516 | 13 | 466 | Normal | Normal |
|  |  | 9 | 851 | 13 | 766 |  |  |
|  |  | X | 1182 | Y |  | Female | Female |
|  |  | X | 517 | Y |  |  |  |
|  |  | 3 | 741 | X | 748 |  |  |
|  |  | 18 | 626 | X | 663 |  |  |
| 7 | 34986 | 11 | 1671 | 21 | 1855 | Normal | Normal |
|  |  | 6 | 2418 | 21 | 2370 |  |  |
|  |  | 10 | 2117 | 18 | 1857 | Normal | Normal |
|  |  | 1 | 1776 | 18 | 1756 |  |  |
|  |  | 11 | 1604 | 13 | 1584 | Normal | Normal |
|  |  | 9 | 1768 | 13 | 1561 |  |  |
|  |  | X | 1162 | Y | 971 | Male | Male |
|  |  | X | 749 | Y | 753 |  |  |
|  |  | 3 | 1941 | X | 942 |  |  |
|  |  | 18 | 1913 | X | 1065 |  |  |
| 8 | 34978 | 11 | 1112 | 21 | 1105 | Normal | Normal |
|  |  | 6 | 887 | 21 | 907 |  |  |
|  |  | 10 | 1268 | 18 | 1046 | Normal | Normal |
|  |  | 1 | 1136 | 18 | 1103 |  |  |
|  |  | 11 | 701 | 13 | 658 | Normal | Normal |
|  |  | 9 | 1903 | 13 | 1546 |  |  |
|  |  | X | 1095 | Y |  | Female | Female |
|  |  | X | 881 | Y |  |  |  |
|  |  | 3 | 614 | X | 606 |  |  |
|  |  | 18 | 899 | X | 883 |  |  |
| 9 | 34731 | 11 | 6168 | 21 | 6027 | Normal | Normal |
|  |  | 6 | 5765 | 21 | 5833 |  |  |
| 10 | 34883 | 11 | 5947 | 21 | 5634 | Normal | Normal |
|  |  | 6 | 6559 | 21 | 6463 |  |  |
| 11 | 35157 | 11 | 983 | 21 | 932 | Normal | Normal |
|  |  | 6 | 1049 | 21 | 1015 |  |  |
|  |  | 10 | 1390 | 18 | 1206 | Normal | Normal |
|  |  | 1 | 928 | 18 | 948 |  |  |
|  |  | 11 | 854 | 13 | 798 | Normal | Normal |
|  |  | 9 | 1395 | 13 | 1192 |  |  |
|  |  | X | 610 | Y | 508 | Male | Male |
|  |  | X | 475 | Y | 498 |  |  |
|  |  | 3 | 872 | X | 425 |  |  |
|  |  | 18 | 988 | X | 500 |  |  |
| 12 | 35125 | 11 | 1954 | 21 | 2146 | Normal | Normal |
|  |  | 6 | 1968 | 21 | 1776 |  |  |
| 13 | 35238 | 10 | 4497 | 18 | 3898 | Normal | Normal |
|  |  | 1 | 1283 | 18 | 1340 |  |  |
|  |  | 11 | 856 | 13 | 805 | Normal | Normal |
|  |  | 9 | 1934 | 13 | 1710 |  |  |
| 14 | 35240 | 11 | 4180 | 21 | 4257 | Normal | Normal |
|  |  | 6 | 4232 | 21 | 4100 |  |  |
| 15 | 35241 | 11 | 4519 | 21 | 4396 | Normal | Normal |
|  |  | 6 | 4613 | 21 | 4137 |  |  |
| 16 | 35645 | 11 | 3589 | 21 | 3660 | Normal | Normal |
|  |  | 6 | 3629 | 21 | 3571 |  |  |
| 17 | 35657 | 11 | 6632 | 21 | 6841 | Normal | Normal |
|  |  | 6 | 8424 | 21 | 8402 |  |  |
| 18 | 35764 | 11 | 3489 | 21 | 3556 | Normal | Normal |
|  |  | 6 | 4241 | 21 | 4138 |  |  |
| 19 | 35765 | 11 | 3874 | 21 | 3982 | Normal | Normal |
|  |  | 6 | 4326 | 21 | 4409 |  |  |
| 20 | 35766 | 11 | 2728 | 21 | 2749 | Normal | Normal |
|  |  | 6 | 3502 | 21 | 3221 |  |  |
| 21 | 35767 | 11 | 3389 | 21 | 3404 | Normal | Normal |
|  |  | 6 | 3848 | 21 | 3948 |  |  |
| 22 | 35643 | 11 | 841 | 21 | 849 | Normal | Normal |
|  |  | 6 | 1287 | 21 | 1188 |  |  |
|  |  | 10 | 1419 | 18 | 1141 | Normal | Normal |
|  |  | 1 | 1468 | 18 | 1489 |  |  |
|  |  | 11 | 998 | 13 | 979 | Normal | Normal |
|  |  | 9 | 1781 | 13 | 1589 |  |  |
|  |  | X | 559 | Y | 488 | Male | Male |
|  |  | X | 681 | Y | 654 |  |  |
|  |  | 3 | 968 | X | 459 |  |  |
|  |  | 18 | 1234 | X | 636 |  |  |
| 23 | 35809 | 11 | 3570 | 21 | 3982 | Normal | Normal |
|  |  | 6 | 4396 | 21 | 4310 |  |  |
|  |  | 10 | 5051 | 18 | 4654 | Normal | Normal |
|  |  | 1 | 3418 | 18 | 3319 |  |  |
|  |  | 11 | 3265 | 13 | 3325 | Normal | Normal |
|  |  | 9 | 3846 | 13 | 3355 |  |  |
|  |  | X | 5983 | Y |  | Female | Female |
|  |  | X | 4438 | Y |  |  |  |
|  |  | 3 | 4851 | X | 4868 |  |  |
|  |  | 18 | 3901 | X | 4291 |  |  |
| 24 | 35821 | 11 | 85 | 21 | 142 | Trisomy | Trisomy |
|  |  | 6 | 118 | 21 | 203 |  |  |
|  |  | 10 | 180 | 18 | 147 | Normal | Normal |
|  |  | 1 | 186 | 18 | 175 |  |  |
|  |  | 11 | 168 | 13 | 148 | Normal | Normal |
|  |  | 9 | 305 | 13 | 260 |  |  |
|  |  | X | 80 | Y | 70 | Male | Male |
|  |  | X | 124 | Y | 138 |  |  |
|  |  | 3 | 159 | X | 83 |  |  |
|  |  | 18 | 211 | X | 106 |  |  |
| 25 | 35822 | 11 | 731 | 21 | 765 | Normal | Trisomy |
|  |  | 6 | 718 | 21 | 900 |  |  |
|  |  | 10 | 1597 | 18 | 1396 | Normal | Normal |
|  |  | 1 | 1208 | 18 | 1214 |  |  |
|  |  | 11 | 909 | 13 | 911 | Normal | Normal |
|  |  | 9 | 1382 | 13 | 1196 |  |  |
|  |  | X | 1223 | Y |  | Female | Female |
|  |  | X | 1372 | Y |  |  |  |
|  |  | 3 | 1066 | X | 998 |  |  |
|  |  | 18 | 1349 | X | 1334 |  |  |
| 26 | 35856 | 11 | 856 | 21 | 844 | Normal | Normal |
|  |  | 6 | 864 | 21 | 792 |  |  |
|  |  | 10 | 1064 | 18 | 893 | Normal | Normal |
|  |  | 1 | 1335 | 18 | 1347 |  |  |
|  |  | 11 | 839 | 13 | 807 | Normal | Normal |
|  |  | 9 | 1569 | 13 | 1270 |  |  |
|  |  | X | 1057 | Y |  | Female | Female |
|  |  | X | 1112 | Y |  |  |  |
|  |  | 3 | 796 | X | 778 |  |  |
|  |  | 18 | 1155 | X | 1144 |  |  |
| 27 | 35857 | 11 | 1540 | 21 | 1711 | Normal | Normal |
|  |  | 6 | 2053 | 21 | 1990 |  |  |
|  |  | 10 | 1998 | 18 | 1863 | Normal | Normal |
|  |  | 1 | 1446 | 18 | 1422 |  |  |
|  |  | 11 | 1242 | 13 | 1209 | Normal | Normal |
|  |  | 9 | 1639 | 13 | 1424 |  |  |
|  |  | X | 2158 | Y |  | Female | Female |
|  |  | X | 1195 | Y |  |  |  |
|  |  | 3 | 1682 | X | 1637 |  |  |
|  |  | 18 | 1620 | X | 1722 |  |  |
| 28 | 35823 | 11 | 5346 | 21 | 7270 | Trisomy | Trisomy |
|  |  | 6 | 2499 | 21 | 4924 |  |  |
| 29 | 36215 | 11 | 5754 | 21 | 5662 | Normal | Normal |
|  |  | 6 | 5631 | 21 | 5674 |  |  |
| 30 | 36216 | 11 | 4341 | 21 | 4251 | Normal | Normal |
|  |  | 6 | 3793 | 21 | 3749 |  |  |
| 31 | 36258 | 11 | 8435 | 21 | 8750 | Normal | Normal |
|  |  | 6 | 8586 | 21 | 8505 |  |  |
| 32 | 35821 | 11 | 385 | 21 | 532 | Trisomy | Trisomy |
|  |  | 6 | 455 | 21 | 661 |  |  |
|  |  | 10 | 593 | 18 | 510 | Normal | Normal |
|  |  | 1 | 432 | 18 | 430 |  |  |
|  |  | 11 | 289 | 13 | 278 | Normal | Normal |
|  |  | 9 | 621 | 13 | 539 |  |  |
|  |  | X | 341 | Y | 259 | Male | Male |
|  |  | X | 161 | Y | 161 |  |  |
|  |  | 3 | 333 | X | 176 |  |  |
|  |  | 18 | 449 | X | 219 |  |  |
| 33 | 36414 | 11 | 1356 | 21 | 1367 | Normal | Normal |
|  |  | 6 | 1557 | 21 | 1593 |  |  |
|  |  | 10 | 1647 | 18 | 1438 | Normal | Normal |
|  |  | 1 | 1557 | 18 | 1574 |  |  |
|  |  | 11 | 1046 | 13 | 1016 | Normal | Normal |
|  |  | 9 | 1682 | 13 | 1402 |  |  |
|  |  | X | 951 | Y | 788 | Male | Male |
|  |  | X | 709 | Y | 718 |  |  |
|  |  | 3 | 1409 | X | 656 |  |  |
|  |  | 18 | 1335 | X | 678 |  |  |
| 34 | 36579 | 11 | 1200 | 21 | 1217 | Normal | Normal |
|  |  | 6 | 1096 | 21 | 1047 |  |  |
|  |  | 10 | 1454 | 18 | 1281 | Normal | Normal |
|  |  | 1 | 1577 | 18 | 1604 |  |  |
|  |  | 11 | 1176 | 13 | 1122 | Normal | Normal |
|  |  | 9 | 1818 | 13 | 1471 |  |  |
|  |  | X | 692 | Y | 596 | Male | Male |
|  |  | X | 749 | Y | 729 |  |  |
|  |  | 3 | 1068 | X | 511 |  |  |
|  |  | 18 | 1446 | X | 720 |  |  |
| 35 | 36772 | 11 | 2034 | 21 | 2334 | Normal | Normal |
|  |  | 6 | 1840 | 21 | 1734 |  |  |
|  |  | 10 | 2213 | 18 | 2062 | Normal | Normal |
|  |  | 1 | 2407 | 18 | 2448 |  |  |
|  |  | 11 | 1069 | 13 | 1070 | Normal | Normal |
|  |  | 9 | 2014 | 13 | 1817 |  |  |
|  |  | X | 2141 | Y |  | Female | Female |
|  |  | X | 1666 | Y |  |  |  |
|  |  | 3 | 1607 | X | 1470 |  |  |
|  |  | 18 | 1968 | X | 1994 |  |  |
| 36 | 36849 | 11 | 2148 | 21 | 3158 | Trisomy | Trisomy |
|  |  | 6 | 2247 | 21 | 3276 |  |  |
|  |  | 10 | 2471 | 18 | 2112 | Normal | Normal |
|  |  | 1 | 1739 | 18 | 1701 |  |  |
|  |  | 11 | 1209 | 13 | 1204 | Normal | Normal |
|  |  | 9 | 1603 | 13 | 1371 |  |  |
|  |  | X | 1205 | Y | 1018 | Male | Male |
|  |  | X | 682 | Y | 683 |  |  |
|  |  | 3 | 2084 | X | 977 |  |  |
|  |  | 18 | 1799 | X | 915 |  |  |
| 37 | 36943 | 11 | 695 | 21 | 705 | Normal | Normal |
|  |  | 6 | 569 | 21 | 580 |  |  |
|  |  | 10 | 660 | 18 | 554 | Normal | Normal |
|  |  | 1 | 847 | 18 | 824 |  |  |
|  |  | 11 | 613 | 13 | 561 | Normal | Normal |
|  |  | 9 | 1091 | 13 | 966 |  |  |
|  |  | X | 684 | Y |  | Female | Female |
|  |  | X | 720 | Y |  |  |  |
|  |  | 3 | 559 | X | 541 |  |  |
|  |  | 18 | 824 | X | 854 |  |  |
| 38 | 36809 | 11 | 4962 | 21 | 5039 | Normal | Normal |
|  |  | 6 | 4719 | 21 | 4567 |  |  |
| 39 | 36810 | 11 | 4509 | 21 | 4363 | Normal | Normal |
|  |  | 6 | 4002 | 21 | 3930 |  |  |
| 40 | 36811 | 11 | 5092 | 21 | 5320 | Normal | Normal |
|  |  | 6 | 4349 | 21 | 4158 |  |  |
| 41 | 13011 | 11 | 667 | 21 | 1169 | Trisomy | Trisomy |
|  |  | 6 | 1442 | 21 | 1913 |  |  |
| 42 | 37012 | 11 | 1455 | 21 | 1483 | Normal | Normal |
|  |  | 6 | 1567 | 21 | 1487 |  |  |
|  |  | 10 | 1822 | 18 | 1565 | Normal | Normal |
|  |  | 1 | 1117 | 18 | 1089 |  |  |
|  |  | 11 | 718 | 13 | 713 | Normal | Normal |
|  |  | 9 | 1379 | 13 | 1189 |  |  |
|  |  | X | 896 | Y | 768 | Male | Male |
|  |  | X | 590 | Y | 569 |  |  |
|  |  | 3 | 1280 | X | 598 |  |  |
|  |  | 18 | 1194 | X | 634 |  |  |
| 43 | 37030 | 11 | 2272 | 21 | 2344 | Normal | Normal |
|  |  | 6 | 2228 | 21 | 2360 |  |  |
|  |  | 10 | 2648 | 18 | 2237 | Normal | Normal |
|  |  | 1 | 2940 | 18 | 2945 |  |  |
|  |  | 11 | 2203 | 13 | 2104 | Normal | Normal |
|  |  | 9 | 3251 | 13 | 2740 |  |  |
|  |  | X | 2821 | Y |  | Female | Female |
|  |  | X | 2484 | Y |  |  |  |
|  |  | 3 | 2175 | X | 2061 |  |  |
|  |  | 18 | 3111 | X | 3195 |  |  |
| 44 | 37243 | 11 | 7733 | 21 | 7663 | Normal | Normal |
|  |  | 6 | 6794 | 21 | 6964 |  |  |
| 45 | 37244 | 11 | 4991 | 21 | 5030 | Normal | Normal |
|  |  | 6 | 5023 | 21 | 5036 |  |  |
| 46 | 37245 | 11 | 4596 | 21 | 4559 | Normal | Normal |
|  |  | 6 | 3833 | 21 | 3806 |  |  |
| 47 | 37246 | 11 | 3169 | 21 | 3207 | Normal | Normal |
|  |  | 6 | 1775 | 21 | 1808 |  |  |
| 48 | 37465 | 11 | 8564 | 21 | 8620 | Normal | Normal |
|  |  | 6 | 8045 | 21 | 7828 |  |  |
| 49 | 37228 | 11 | 2039 | 21 | 2008 | Normal | Normal |
|  |  | 6 | 2621 | 21 | 2445 |  |  |
|  |  | 10 | 3115 | 18 | 2758 | Normal | Normal |
|  |  | 1 | 2249 | 18 | 2307 |  |  |
|  |  | 11 | 1673 | 13 | 1562 | Normal | Normal |
|  |  | 9 | 2332 | 13 | 2076 |  |  |
|  |  | X | 1266 | Y | 1048 | Male | Male |
|  |  | X | 993 | Y | 1018 |  |  |
|  |  | 3 | 2219 | X | 1085 |  |  |
|  |  | 18 | 2273 | X | 1149 |  |  |
| 50 | 37239 | 11 | 965 | 21 | 966 | Normal | Normal |
|  |  | 6 | 1012 | 21 | 1049 |  |  |
|  |  | 10 | 1344 | 18 | 1130 | Normal | Normal |
|  |  | 1 | 1306 | 18 | 1331 |  |  |
|  |  | 11 | 941 | 13 | 913 | Normal | Normal |
|  |  | 9 | 1538 | 13 | 1338 |  |  |
|  |  | X | 982 | Y |  | Female | Female |
|  |  | X | 1206 | Y |  |  |  |
|  |  | 3 | 901 | X | 849 |  |  |
|  |  | 18 | 1229 | X | 1257 |  |  |
| 51 | 37390 | 11 | 3602 | 21 | 3538 | Normal | Normal |
|  |  | 6 | 3798 | 21 | 3561 |  |  |
|  |  | 10 | 4820 | 18 | 4295 | Normal | Normal |
|  |  | 1 | 3201 | 18 | 3204 |  |  |
|  |  | 11 | 2396 | 13 | 2394 | Normal | Normal |
|  |  | 9 | 3159 | 13 | 2657 |  |  |
|  |  | X | 4225 | Y |  | Female | Female |
|  |  | X | 3172 | Y |  |  |  |
|  |  | 3 | 3842 | X | 3889 |  |  |
|  |  | 18 | 3505 | X | 3754 |  |  |
| 52 | 37450 | 11 | 3076 | 21 | 3241 | Normal | Normal |
|  |  | 6 | 3927 | 21 | 3713 |  |  |
|  |  | 10 | 4223 | 18 | 3746 | Normal | Normal |
|  |  | 1 | 4340 | 18 | 4264 |  |  |
|  |  | 11 | 3288 | 13 | 3165 | Normal | Normal |
|  |  | 9 | 4230 | 13 | 3709 |  |  |
|  |  | X | 4431 | Y |  | Female | Female |
|  |  | X | 3688 | Y |  |  |  |
|  |  | 3 | 3725 | X | 3765 |  |  |
|  |  | 18 | 4047 | X | 4277 |  |  |
| 53 | 37529 | 11 | 3793 | 21 | 3911 | Normal | Normal |
|  |  | 6 | 3103 | 21 | 3000 |  |  |
| 54 | 37531 | 11 | 2966 | 21 | 2930 | Normal | Normal |
|  |  | 6 | 3998 | 21 | 4070 |  |  |
| 55 | 37532 | 11 | 4187 | 21 | 4139 | Normal | Normal |
|  |  | 6 | 4400 | 21 | 4332 |  |  |
| 56 | 37622 | 11 | 4860 | 21 | 4994 | Normal | Normal |
|  |  | 6 | 6140 | 21 | 6295 |  |  |
| 57 | 37530 | 10 | 6022 | 18 | 4887 | Normal | Normal |
|  |  | 1 | 4833 | 18 | 4701 |  |  |
| 58 | 37688 | 11 | 1343 | 21 | 1512 | Normal | Normal |
|  |  | 6 | 1814 | 21 | 1724 |  |  |
|  |  | 10 | 1984 | 18 | 1843 | Normal | Normal |
|  |  | 1 | 2387 | 18 | 2296 |  |  |
|  |  | 11 | 2030 | 13 | 1965 | Normal | Normal |
|  |  | 9 | 2374 | 13 | 2116 |  |  |
|  |  | X | 1078 | Y | 912 | Male | Male |
|  |  | X | 1156 | Y | 1142 |  |  |
|  |  | 3 | 1552 | X | 815 |  |  |
|  |  | 18 | 2472 | X | 1395 |  |  |
| 59 | 37914 | 11 | 2626 | 21 | 2745 | Normal | Normal |
|  |  | 6 | 2916 | 21 | 2788 |  |  |
| 60 | 37915 | 11 | 3696 | 21 | 5248 | Trisomy | Trisomy |
|  |  | 6 | 3582 | 21 | 5329 |  |  |
| 61 | 37936 | 11 | 818 | 21 | 1014 | Trisomy | Trisomy |
|  |  | 6 | 664 | 21 | 836 |  |  |
|  |  | 10 | 1013 | 18 | 906 | Normal | Normal |
|  |  | 1 | 1216 | 18 | 1283 |  |  |
|  |  | 11 | 1027 | 13 | 999 | Normal | Normal |
|  |  | 9 | 1426 | 13 | 1309 |  |  |
|  |  | X | 916 | Y |  | Female | Female |
|  |  | X | 1008 | Y |  |  |  |
|  |  | 3 | 754 | X | 741 |  |  |
|  |  | 18 | 1373 | X | 1422 |  |  |
| 62 | 37951 | 11 | 727 | 21 | 828 | Normal | Normal |
|  |  | 6 | 1015 | 21 | 981 |  |  |
|  |  | 10 | 1174 | **18** | 1559 | Trisomy | Trisomy |
|  |  | 1 | 1028 | **18** | 1524 |  |  |
|  |  | 11 | 1050 | 13 | 1008 | Normal | Normal |
|  |  | 9 | 1093 | 13 | 1019 |  |  |
|  |  | X | 1209 | Y |  | Female | Female |
|  |  | X | 1073 | Y |  |  |  |
|  |  | 3 | 1004 | X | 961 |  |  |
|  |  | 18 | 1835 | X | 1798 |  |  |
| 63 | 37978 | 11 | 865 | 21 | 923 | Normal | Normal |
|  |  | 6 | 968 | 21 | 990 |  |  |
|  |  | 10 | 1103 | 18 | 917 | Normal | Normal |
|  |  | 1 | 1045 | 18 | 1067 |  |  |
|  |  | 11 | 931 | 13 | 917 | Normal | Normal |
|  |  | 9 | 1667 | 13 | 1422 |  |  |
|  |  | X | 505 | Y | 445 | Male | Male |
|  |  | X | 526 | Y | 527 |  |  |
|  |  | 3 | 736 | X | 376 |  |  |
|  |  | 18 | 1361 | X | 679 |  |  |
| 64 | 38050 | 11 | 437 | 21 | 462 | Normal | Normal |
|  |  | 6 | 498 | 21 | 551 |  |  |
|  |  | 10 | 693 | 18 | 536 | Normal | Normal |
|  |  | 1 | 608 | 18 | 573 |  |  |
|  |  | 11 | 410 | 13 | 390 | Normal | Normal |
|  |  | 9 | 1055 | 13 | 847 |  |  |
|  |  | X | 560 | Y |  | Female | Female |
|  |  | X | 493 | Y |  |  |  |
|  |  | 3 | 387 | X | 376 |  |  |
|  |  | 18 | 544 | X | 513 |  |  |
| 65 | 38056 | 10 | 4059 | 18 | 3475 | Normal | Normal |
|  |  | 1 | 3085 | 18 | 3087 |  |  |
| 66 | 38244 | 11 | 2194 | 21 | 2242 | Normal | Normal |
|  |  | 6 | 2391 | 21 | 2016 |  |  |
|  |  | 10 | 3380 | 18 | 2689 | Normal | Normal |
|  |  | 1 | 2602 | 18 | 2537 |  |  |
|  |  | 11 | 2447 | 13 | 2415 | Normal | Normal |
|  |  | 9 | 2244 | 13 | 1887 |  |  |
|  |  | X | 1346 | Y | 1278 | Male | Male |
|  |  | X | 1260 | Y | 1320 |  |  |
|  |  | 3 | 2331 | X | 1136 |  |  |
|  |  | 18 | 2408 | X | 1377 |  |  |
| 67 | 38245 | 11 | 1836 | 21 | 1838 | Normal | Normal |
|  |  | 6 | 1682 | 21 | 1517 |  |  |
|  |  | 10 | 2492 | 18 | 2079 | Normal | Normal |
|  |  | 1 | 2997 | 18 | 2997 |  |  |
|  |  | 11 | 1938 | 13 | 1829 | Normal | Normal |
|  |  | 9 | 3154 | 13 | 2731 |  |  |
|  |  | X | 2096 | Y |  | Female | Female |
|  |  | X | 2818 | Y |  |  |  |
|  |  | 3 | 1554 | X | 1496 |  |  |
|  |  | 18 | 2843 | X | 2775 |  |  |
| 68 | 38247 | 11 | 1468 | 21 | 1426 | Normal | Normal |
|  |  | 6 | 1440 | 21 | 1398 |  |  |
|  |  | 10 | 1867 | 18 | 1598 | Normal | Normal |
|  |  | 1 | 1179 | 18 | 1165 |  |  |
|  |  | 11 | 795 | 13 | 719 | Normal | Normal |
|  |  | 9 | 1415 | 13 | 1144 |  |  |
|  |  | X | 801 | Y | 695 | Male | Male |
|  |  | X | 531 | Y | 572 |  |  |
|  |  | 3 | 1107 | X | 532 |  |  |
|  |  | 18 | 1056 | X | 563 |  |  |
| 69 | 38248 | 11 | 1276 | 21 | 1228 | Normal | Normal |
|  |  | 6 | 1097 | 21 | 1052 |  |  |
|  |  | 10 | 1544 | 18 | 1267 | Normal | Normal |
|  |  | 1 | 1855 | 18 | 1964 |  |  |
|  |  | 11 | 1245 | 13 | 1167 | Normal | Normal |
|  |  | 9 | 2193 | 13 | 1924 |  |  |
|  |  | X | 1456 | Y |  | Female | Female |
|  |  | X | 1701 | Y |  |  |  |
|  |  | 3 | 1016 | X | 925 |  |  |
|  |  | 18 | 1589 | X | 1605 |  |  |
| 70 | 38275 | 11 | 3048 | 21 | 3012 | Normal | Normal |
|  |  | 6 | 3162 | 21 | 3007 |  |  |
|  |  | 10 | 3501 | 18 | 2869 | Normal | Normal |
|  |  | 1 | 2955 | 18 | 2941 |  |  |
|  |  | 11 | 2366 | 13 | 2467 | Normal | Normal |
|  |  | 9 | 2743 | 13 | 2287 |  |  |
|  |  | X | 3212 | Y |  | Female | Female |
|  |  | X | 2436 | Y |  |  |  |
|  |  | 3 | 2439 | X | 2243 |  |  |
|  |  | 18 | 2786 | X | 2668 |  |  |
| 71 | 38565 | 11 | 525 | 21 | 596 | Normal | Normal |
|  |  | 6 | 757 | 21 | 806 |  |  |
|  |  | 10 | 747 | 18 | 670 | Normal | Normal |
|  |  | 1 | 1116 | 18 | 1147 |  |  |
|  |  | 11 | 876 | 13 | 897 | Normal | Normal |
|  |  | 9 | 1302 | 13 | 1178 |  |  |
|  |  | X | 435 | Y | 392 | Male | Male |
|  |  | X | 493 | Y | 486 |  |  |
|  |  | 3 | 658 | X | 328 |  |  |
|  |  | 18 | 1300 | X | 692 |  |  |
| 72 | 38623 | 11 | 667 | 21 | 642 | Normal | Normal |
|  |  | 6 | 494 | 21 | 452 |  |  |
|  |  | 10 | 788 | 18 | 591 | Normal | Normal |
|  |  | 1 | 514 | 18 | 554 |  |  |
|  |  | 11 | 325 | 13 | 307 | Normal | Normal |
|  |  | 9 | 769 | 13 | 648 |  |  |
|  |  | X | 291 | Y | 231 | Male | Male |
|  |  | X | 200 | Y | 201 |  |  |
|  |  | 3 | 352 | X | 153 |  |  |
|  |  | 18 | 356 | X | 192 |  |  |
| 73 | 38627 | 11 | 2108 | 21 | 2026 | Normal | Normal |
|  |  | 6 | 1907 | 21 | 1843 |  |  |
|  |  | 10 | 2483 | 18 | 2076 | Normal | Normal |
|  |  | 1 | 2673 | 18 | 2690 |  |  |
|  |  | 11 | 1628 | 13 | 1480 | Normal | Normal |
|  |  | 9 | 2896 | 13 | 2553 |  |  |
|  |  | X | 1077 | Y | 893 | Male | Male |
|  |  | X | 1052 | Y | 1074 |  |  |
|  |  | 3 | 1511 | X | 703 |  |  |
|  |  | 18 | 2219 | X | 1128 |  |  |
| 74 | 38630 | 11 | 4568 | 21 | 4273 | Normal | Normal |
|  |  | 6 | 3299 | 21 | 3151 |  |  |
|  |  | 10 | 4759 | 18 | 3546 | Normal | Normal |
|  |  | 1 | 3324 | 18 | 3462 |  |  |
|  |  | 11 | 2637 | 13 | 2520 | Normal | Normal |
|  |  | 9 | 3739 | 13 | 3313 |  |  |
|  |  | X | 3747 | Y |  | Female | Female |
|  |  | X | 3566 | Y |  |  |  |
|  |  | 3 | 2595 | X | 2564 |  |  |
|  |  | 18 | 3208 | X | 3300 |  |  |
| 75 | 35639 | 11 | 1935 | 21 | 1856 | Normal | Normal |
|  |  | 6 | 1518 | 21 | 1664 |  |  |
|  |  | 10 | 2106 | 18 | 1700 | Normal | Normal |
|  |  | 1 | 2893 | 18 | 2951 |  |  |
|  |  | 11 | 2113 | 13 | 1979 | Normal | Normal |
|  |  | 9 | 3049 | 13 | 2676 |  |  |
|  |  | X | 989 | Y | 855 | Male | Male |
|  |  | X | 1315 | Y | 1425 |  |  |
|  |  | 3 | 1262 | X | 625 |  |  |
|  |  | 18 | 2392 | X | 1224 |  |  |
| 76 | 38641 | 11 | 2204 | 21 | 2496 | Normal | Normal |
|  |  | 6 | 2966 | 21 | 2821 |  |  |
|  |  | 10 | 3064 | 18 | 2764 | Normal | Normal |
|  |  | 1 | 2047 | 18 | 2030 |  |  |
|  |  | 11 | 1703 | 13 | 1628 | Normal | Normal |
|  |  | 9 | 2008 | 13 | 1664 |  |  |
|  |  | X | 3221 | Y |  | Female | Female |
|  |  | X | 1778 | Y |  |  |  |
|  |  | 3 | 2368 | X | 2229 |  |  |
|  |  | 18 | 2220 | X | 2309 |  |  |
| 77 | 38665 | 11 | 2300 | 21 | 2361 | Normal | Normal |
|  |  | 6 | 2421 | 21 | 2425 |  |  |
|  |  | 10 | 2647 | 18 | 2118 | Normal | Normal |
|  |  | 1 | 2924 | 18 | 3060 |  |  |
|  |  | 11 | 2283 | 13 | 2118 | Normal | Normal |
|  |  | 9 | 2952 | 13 | 2689 |  |  |
|  |  | X | 1332 | Y | 1142 | Male | Male |
|  |  | X | 1231 | Y | 1233 |  |  |
|  |  | 3 | 1682 | X | 787 |  |  |
|  |  | 18 | 2862 | X | 1453 |  |  |
| 78 | 38054 | 11 | 2840 | 21 | 2918 | Normal | Normal |
|  |  | 6 | 3364 | 21 | 3224 |  |  |
| 79 | 35172 | 11 | 1847 | 21 | 1860 | Normal | Normal |
|  |  | 6 | 2259 | 21 | 2097 |  |  |
|  |  | 10 | 2814 | 18 | 2408 | Normal | Normal |
|  |  | 1 | 2154 | 18 | 2127 |  |  |
|  |  | 11 | 1626 | 13 | 1533 | Normal | Normal |
|  |  | 9 | 2577 | 13 | 2177 |  |  |
|  |  | X | 1130 | Y | 1000 | Male | Male |
|  |  | X | 1021 | Y | 990 |  |  |
|  |  | 3 | 1783 | X | 808 |  |  |
|  |  | 18 | 1922 | X | 1005 |  |  |
| 80 | 38720 | 11 | 1968 | 21 | 1867 | Normal | Normal |
|  |  | 6 | 1152 | 21 | 1129 |  |  |
|  |  | 10 | 2008 | 18 | 1556 | Normal | Normal |
|  |  | 1 | 2166 | 18 | 2019 |  |  |
|  |  | 11 | 1186 | 13 | 1124 | Normal | Normal |
|  |  | 9 | 3212 | 13 | 2499 |  |  |
|  |  | X | 1349 | Y |  | Female | Female |
|  |  | X | 1638 | Y |  |  |  |
|  |  | 3 | 954 | X | 901 |  |  |
|  |  | 18 | 1643 | X | 1627 |  |  |
| 81 | 38780 | 11 | 2008 | 21 | 1986 | Normal | Normal |
|  |  | 6 | 1649 | 21 | 1615 |  |  |
|  |  | 10 | 2459 | 18 | 1966 | Normal | Normal |
|  |  | 1 | 1580 | 18 | 1548 |  |  |
|  |  | 11 | 958 | 13 | 917 | Normal | Normal |
|  |  | 9 | 2262 | 13 | 1803 |  |  |
|  |  | X | 1882 | Y |  | Female | Female |
|  |  | X | 1415 | Y |  |  |  |
|  |  | 3 | 1294 | X | 1220 |  |  |
|  |  | 18 | 1366 | X | 1321 |  |  |
| 82 | 38733 | 11 | 6412 | 21 | 6263 | Normal | Normal |
|  |  | 6 | 6032 | 21 | 5835 |  |  |
| 83 | 38734 | 11 | 6214 | 21 | 6072 | Normal | Normal |
|  |  | 6 | 4957 | 21 | 4914 |  |  |
| 84 | 38911 | 11 | 4539 | 21 | 4616 | Normal | Normal |
|  |  | 6 | 4517 | 21 | 4521 |  |  |
|  |  | 10 | 7273 | 18 | 6827 | Normal | Normal |
|  |  | 1 | 4625 | 18 | 4744 |  |  |
|  |  | 11 | 4909 | 13 | 4494 | Normal | Normal |
|  |  | 9 | 4953 | 13 | 4527 |  |  |
|  |  | X | 6560 | Y |  | Female | Female |
|  |  | X | 6078 | Y |  |  |  |
|  |  | 3 | 4540 | X | 4133 |  |  |
|  |  | 18 | 6066 | X | 6522 |  |  |
| 85 | 38918 | 11 | 1408 | 21 | 1362 | Normal | Normal |
|  |  | 6 | 1287 | 21 | 1232 |  |  |
|  |  | 10 | 1747 | 18 | 1478 | Normal | Normal |
|  |  | 1 | 1743 | 18 | 1847 |  |  |
|  |  | 11 | 1252 | 13 | 1117 | Normal | Normal |
|  |  | 9 | 2585 | 13 | 2103 |  |  |
|  |  | X | 748 | Y | 602 | Male | Male |
|  |  | X | 736 | Y | 759 |  |  |
|  |  | 3 | 892 | X | 425 |  |  |
|  |  | 18 | 1653 | X | 804 |  |  |
| 86 | 38924 | 11 | 2439 | 21 | 2486 | Normal | Normal |
|  |  | 6 | 2140 | 21 | 2201 |  |  |
|  |  | 10 | 2920 | 18 | 2547 | Normal | Normal |
|  |  | 1 | 1952 | 18 | 1882 |  |  |
|  |  | 11 | 1551 | 13 | 1443 | Normal | Normal |
|  |  | 9 | 2758 | 13 | 2297 |  |  |
|  |  | X | 2567 | Y |  | Female | Female |
|  |  | X | 1684 | Y |  |  |  |
|  |  | 3 | 1689 | X | 1663 |  |  |
|  |  | 18 | 1825 | X | 1938 |  |  |
| 87 | 38986 | 11 | 1986 | 21 | 1942 | Normal | Normal |
|  |  | 6 | 1873 | 21 | 1750 |  |  |
|  |  | 10 | 2389 | 18 | 2057 | Normal | Normal |
|  |  | 1 | 2874 | 18 | 2879 |  |  |
|  |  | 11 | 2022 | 13 | 1907 | Normal | Normal |
|  |  | 9 | 3600 | 13 | 2960 |  |  |
|  |  | X | 1091 | Y | 956 | Male | Male |
|  |  | X | 1188 | Y | 1187 |  |  |
|  |  | 3 | 1465 | X | 699 |  |  |
|  |  | 18 | 2517 | X | 1256 |  |  |
| 88 | 38988 | 11 | 2286 | 21 | 2646 | Normal | Normal |
|  |  | 6 | 2506 | 21 | 2492 |  |  |
|  |  | 10 | 2919 | 18 | 2536 | Normal | Normal |
|  |  | 1 | 2491 | 18 | 2580 |  |  |
|  |  | 11 | 2122 | 13 | 1969 | Normal | Normal |
|  |  | 9 | 2980 | 13 | 2684 |  |  |
|  |  | X | 1560 | Y | 1292 | Male | Male |
|  |  | X | 1122 | Y | 1121 |  |  |
|  |  | 3 | 2039 | X | 974 |  |  |
|  |  | 18 | 2408 | X | 1369 |  |  |
| 89 | 38992 | 11 | 1193 | 21 | 1223 | Normal | Normal |
|  |  | 6 | 1195 | 21 | 1186 |  |  |
|  |  | 10 | 1661 | 18 | 1359 | Normal | Normal |
|  |  | 1 | 1867 | 18 | 1981 |  |  |
|  |  | 11 | 1350 | 13 | 1357 | Normal | Normal |
|  |  | 9 | 2501 | 13 | 2029 |  |  |
|  |  | X | 733 | Y | 582 | Male | Male |
|  |  | X | 757 | Y | 761 |  |  |
|  |  | 3 | 997 | X | 489 |  |  |
|  |  | 18 | 1495 | X | 764 |  |  |
| 90 | 39018 | 11 | 3841 | 21 | 3395 | Normal | Normal |
|  |  | 6 | 3294 | 21 | 3035 |  |  |
|  |  | 10 | 4664 | 18 | 3438 | Normal | Normal |
|  |  | 1 | 2536 | 18 | 2806 |  |  |
|  |  | 11 | 2163 | 13 | 1953 | Normal | Normal |
|  |  | 9 | 2673 | 13 | 2291 |  |  |
|  |  | X | 1804 | Y | 1709 | Male | Male |
|  |  | X | 1367 | Y | 1612 |  |  |
|  |  | 3 | 2745 | X | 1427 |  |  |
|  |  | 18 | 2372 | X | 1246 |  |  |
| 91 | 39039 | 11 | 4501 | 21 | 4326 | Normal | Normal |
|  |  | 6 | 3732 | 21 | 3741 |  |  |
|  |  | 10 | 5530 | 18 | 4486 | Normal | Normal |
|  |  | 1 | 5342 | 18 | 5431 |  |  |
|  |  | 11 | 3663 | 13 | 3696 | Normal | Normal |
|  |  | 9 | 5308 | 13 | 4419 |  |  |
|  |  | X | 4577 | Y |  | Female | Female |
|  |  | X | 4649 | Y |  |  |  |
|  |  | 3 | 3517 | X | 3334 |  |  |
|  |  | 18 | 4391 | X | 4538 |  |  |
| 92 | 39040 | 11 | 2508 | 21 | 2486 | Normal | Normal |
|  |  | 6 | 2414 | 21 | 2418 |  |  |
|  |  | 10 | 3397 | 18 | 2696 | Normal | Normal |
|  |  | 1 | 2404 | 18 | 2354 |  |  |
|  |  | 11 | 1017 | 13 | 919 | Normal | Normal |
|  |  | 9 | 1716 | 13 | 1379 |  |  |
|  |  | X | 1587 | Y | 1320 | Male | Male |
|  |  | X | 1066 | Y | 1139 |  |  |
|  |  | 3 | 2024 | X | 937 |  |  |
|  |  | 18 | 2172 | X | 1159 |  |  |
| 93 | 38931 | 11 | 9015 | 21 | 12621 | Trisomy | Trisomy |
|  |  | 6 | 7358 | 21 | 9174 |  |  |
| 94 | 38932 | 11 | 7640 | 21 | 7458 | Normal | Normal |
|  |  | 6 | 6653 | 21 | 7091 |  |  |
| 95 | 39118 | 11 | 1549 | 21 | 1568 | Normal | Normal |
|  |  | 6 | 1427 | 21 | 1354 |  |  |
|  |  | 10 | 2311 | 18 | 1772 | Normal | Normal |
|  |  | 1 | 1889 | 18 | 1763 |  |  |
|  |  | 11 | 2021 | 13 | 2019 | Normal | Normal |
|  |  | 9 | 2266 | 13 | 1876 |  |  |
|  |  | X | 725 | Y | 673 | Male | Male |
|  |  | X | 911 | Y | 900 |  |  |
|  |  | 3 | 1325 | X | 640 |  |  |
|  |  | 18 | 1685 | X | 842 |  |  |
| 96 | 39119 | 11 | 1092 | 21 | 1003 | Normal | Normal |
|  |  | 6 | 800 | 21 | 703 |  |  |
|  |  | 10 | 1975 | 18 | 1631 | Normal | Normal |
|  |  | 1 | 566 | 18 | 581 |  |  |
|  |  | 11 | 804 | 13 | 728 | Normal | Normal |
|  |  | 9 | 1016 | 13 | 736 |  |  |
|  |  | X | 636 | Y | 552 | Male | Male |
|  |  | X | 422 | Y | 427 |  |  |
|  |  | 3 | 1198 | X | 403 |  |  |
|  |  | 18 | 716 | X | 365 |  |  |
| 97 | 39062 | 11 | 851 | 21 | 757 | Normal | Normal |
|  |  | 6 | 523 | 21 | 432 |  |  |
|  |  | 10 | 1535 | **18** | 2133 | Trisomy | Trisomy |
|  |  | 1 | 771 | **18** | 1005 |  |  |
|  |  | 11 | 843 | 13 | 628 | Normal | Normal |
|  |  | 9 | 1066 | 13 | 925 |  |  |
|  |  | X | 873 | Y |  | Female | Female |
|  |  | X | 1373 | Y |  |  |  |
|  |  | 3 | 330 | X | 406 |  |  |
|  |  | 18 | 720 | X | 642 |  |  |
| 98 | 39158 | 11 | 1939 | 21 | 1912 | Normal | Normal |
|  |  | 6 | 2080 | 21 | 2015 |  |  |
|  |  | 10 | 2270 | 18 | 2000 | Normal | Normal |
|  |  | 1 | 2239 | 18 | 2241 |  |  |
|  |  | 11 | 1578 | 13 | 1537 | Normal | Normal |
|  |  | 9 | 2400 | 13 | 2089 |  |  |
|  |  | X | 1037 | Y | 889 | Male | Male |
|  |  | X | 972 | Y | 986 |  |  |
|  |  | 3 | 1635 | X | 817 |  |  |
|  |  | 18 | 2234 | X | 1114 |  |  |
| 99 | 39302 | 11 | 2352 | 21 | 2304 | Normal | Normal |
|  |  | 6 | 2115 | 21 | 2051 |  |  |
|  |  | 10 | 2676 | 18 | 2227 | Normal | Normal |
|  |  | 1 | 2709 | 18 | 2807 |  |  |
|  |  | 11 | 2344 | 13 | 2365 | Normal | Normal |
|  |  | 9 | 3122 | 13 | 2710 |  |  |
|  |  | X | 2722 | Y |  | Female | Female |
|  |  | X | 2970 | Y |  |  |  |
|  |  | 3 | 1908 | X | 1779 |  |  |
|  |  | 18 | 2819 | X | 3062 |  |  |
| 100 | 39375 | 11 | 1211 | 21 | 1220 | Normal | Normal |
|  |  | 6 | 1372 | 21 | 1319 |  |  |
|  |  | 10 | 1705 | 18 | 1477 | Normal | Normal |
|  |  | 1 | 1147 | 18 | 1238 |  |  |
|  |  | 11 | 1181 | 13 | 1136 | Normal | Normal |
|  |  | 9 | 1460 | 13 | 1352 |  |  |
|  |  | X | 1530 | Y |  | Female | Female |
|  |  | X | 1197 | Y |  |  |  |
|  |  | 3 | 1221 | X | 1114 |  |  |
|  |  | 18 | 1498 | X | 1515 |  |  |
| 101 | 39386 | 11 | 5392 | 21 | 5231 | Normal | Normal |
|  |  | 6 | 4434 | 21 | 4495 |  |  |
|  |  | 10 | 6726 | 18 | 5352 | Normal | Normal |
|  |  | 1 | 5608 | 18 | 5577 |  |  |
|  |  | 11 | 4632 | 13 | 4881 | Normal | Normal |
|  |  | 9 | 6066 | 13 | 5053 |  |  |
|  |  | X | 6057 | Y |  | Female | Female |
|  |  | X | 6567 | Y |  |  |  |
|  |  | 3 | 4754 | X | 4794 |  |  |
|  |  | 18 | 6061 | X | 6504 |  |  |
| 102 | 39508 | 11 | 3692 | 21 | 3469 | Normal | Normal |
|  |  | 6 | 4214 | 21 | 4250 |  |  |
| 103 | 39510 | 11 | 4227 | 21 | 4518 | Normal | Normal |
|  |  | 6 | 5507 | 21 | 5392 |  |  |
| 104 | 39511 | 11 | 4682 | 21 | 4816 | Normal | Normal |
|  |  | 6 | 4969 | 21 | 4742 |  |  |
| 105 | 39663 | 11 | 3312 | 21 | 3340 | Normal | Normal |
|  |  | 6 | 3577 | 21 | 3605 |  |  |
|  |  | 10 | 4140 | 18 | 3752 | Normal | Normal |
|  |  | 1 | 3153 | 18 | 3268 |  |  |
|  |  | 11 | 2493 | 13 | 2437 | Normal | Normal |
|  |  | 9 | 3255 | 13 | 2631 |  |  |
|  |  | X | 2139 | Y | 1918 | Male | Male |
|  |  | X | 1623 | Y | 1580 |  |  |
|  |  | 3 | 3439 | X | 1655 |  |  |
|  |  | 18 | 3136 | X | 1662 |  |  |
| 106 | 39695 | 11 | 1969 | 21 | 2044 | Normal | Normal |
|  |  | 6 | 1992 | 21 | 1925 |  |  |
|  |  | 10 | 2684 | 18 | 2230 | Normal | Normal |
|  |  | 1 | 2568 | 18 | 2551 |  |  |
|  |  | 11 | 1739 | 13 | 1649 | Normal | Normal |
|  |  | 9 | 2892 | 13 | 2358 |  |  |
|  |  | X | 2202 | Y |  | Female | Female |
|  |  | X | 2040 | Y |  |  |  |
|  |  | 3 | 1506 | X | 1393 |  |  |
|  |  | 18 | 2380 | X | 2511 |  |  |
| 107 | 39696 | 11 | 2186 | 21 | 2145 | Normal | Normal |
|  |  | 6 | 2194 | 21 | 2055 |  |  |
|  |  | 10 | 2525 | 18 | 2262 | Normal | Normal |
|  |  | 1 | 1789 | 18 | 1755 |  |  |
|  |  | 11 | 1340 | 13 | 1289 | Normal | Normal |
|  |  | 9 | 2209 | 13 | 1866 |  |  |
|  |  | X | 2711 | Y |  | Female | Female |
|  |  | X | 1599 | Y |  |  |  |
|  |  | 3 | 2042 | X | 1952 |  |  |
|  |  | 18 | 1841 | X | 1911 |  |  |
| 108 | 39697 | 11 | 2493 | 21 | 3390 | Trisomy | Trisomy |
|  |  | 6 | 2591 | 21 | 3701 |  |  |
|  |  | 10 | 3418 | 18 | 3134 | Normal | Normal |
|  |  | 1 | 2880 | 18 | 2906 |  |  |
|  |  | 11 | 2350 | 13 | 2246 | Normal | Normal |
|  |  | 9 | 3173 | 13 | 2798 |  |  |
|  |  | X | 2992 | Y |  | Female | Female |
|  |  | X | 2782 | Y |  |  |  |
|  |  | 3 | 2719 | X | 2565 |  |  |
|  |  | 18 | 3236 | X | 3518 |  |  |
| 109 | 39959 | 11 | 4191 | 21 | 6523 | Trisomy | Trisomy |
|  |  | 6 | 4414 | 21 | 6104 |  |  |
|  |  | 10 | 5245 | 18 | 4513 | Normal | Normal |
|  |  | 1 | 4406 | 18 | 4685 |  |  |
|  |  | 11 | 4056 | 13 | 4092 | Normal | Normal |
|  |  | 9 | 5080 | 13 | 4294 |  |  |
|  |  | X | 5408 | Y |  | Female | Female |
|  |  | X | 5166 | Y |  |  |  |
|  |  | 3 | 3769 | X | 3549 |  |  |
|  |  | 18 | 4612 | X | 4788 |  |  |
| 110 | 39857 | 11 | 2232 | 21 | 2152 | Normal | Normal |
|  |  | 6 | 2013 | 21 | 1915 |  |  |
|  |  | 10 | 2490 | 18 | 2167 | Normal | Normal |
|  |  | 1 | 2358 | 18 | 2331 |  |  |
|  |  | 11 | 1612 | 13 | 1510 | Normal | Normal |
|  |  | 9 | 2786 | 13 | 2310 |  |  |
|  |  | X | 2228 | Y |  | Female | Female |
|  |  | X | 1840 | Y |  |  |  |
|  |  | 3 | 1555 | X | 1390 |  |  |
|  |  | 18 | 2135 | X | 2041 |  |  |
| 111 | 39649 | 11 | 1305 | 21 | 1149 | Normal | Normal |
|  |  | 6 | 577 | 21 | 553 |  |  |
|  |  | 10 | 1819 | **18** | 2346 | Trisomy | Trisomy |
|  |  | 1 | 1022 | **18** | 1335 |  |  |
|  |  | 11 | 512 | 13 | 467 | Normal | Normal |
|  |  | 9 | 1287 | 13 | 1027 |  |  |
|  |  | X | 1081 | Y |  | Female | Female |
|  |  | X | 1126 | Y |  |  |  |
|  |  | 3 | 732 | X | 694 |  |  |
|  |  | 18 | 997 | X | 852 |  |  |
| 112 | 39654 | 11 | 4081 | 21 | 3812 | Normal | Normal |
|  |  | 6 | 3573 | 21 | 3532 |  |  |
|  |  | 10 | 5341 | 18 | 3875 | Normal | Normal |
|  |  | 1 | 5213 | 18 | 5002 |  |  |
|  |  | 11 | 3883 | 13 | 3914 | Normal | Normal |
|  |  | 9 | 5272 | 13 | 3953 |  |  |
|  |  | X | 4091 | Y |  | Female | Female |
|  |  | X | 4748 | Y |  |  |  |
|  |  | 3 | 2714 | X | 2515 |  |  |
|  |  | 18 | 4257 | X | 4385 |  |  |
| 113 | 39855 | 11 | 2648 | 21 | 2775 | Normal | Normal |
|  |  | 6 | 2776 | 21 | 2744 |  |  |
|  |  | 10 | 3422 | 18 | 2995 | Normal | Normal |
|  |  | 1 | 2026 | 18 | 2079 |  |  |
|  |  | 11 | 1670 | 13 | 1630 | Normal | Normal |
|  |  | 9 | 2536 | 13 | 2112 |  |  |
|  |  | X | 1573 | Y | 1352 | Male | Male |
|  |  | X | 1110 | Y | 1127 |  |  |
|  |  | 3 | 2667 | X | 1307 |  |  |
|  |  | 18 | 2064 | X | 1057 |  |  |
| 114 | 39863 | 11 | 3240 | 21 | 3252 | Normal | Normal |
|  |  | 6 | 2383 | 21 | 2663 |  |  |
|  |  | 10 | 3317 | 18 | 3006 | Normal | Normal |
|  |  | 1 | 2599 | 18 | 2570 |  |  |
|  |  | 11 | 1909 | 13 | 1909 | Normal | Normal |
|  |  | 9 | 2757 | 13 | 2460 |  |  |
|  |  | X | 3320 | Y |  | Female | Female |
|  |  | X | 2316 | Y |  |  |  |
|  |  | 3 | 2356 | X | 2182 |  |  |
|  |  | 18 | 2331 | X | 2431 |  |  |
| 115 | 39868 | 11 | 6225 | 21 | 5801 | Normal | Normal |
|  |  | 6 | 4946 | 21 | 5190 |  |  |
|  |  | 10 | 6505 | 18 | 5273 | Normal | Normal |
|  |  | 1 | 6913 | 18 | 6676 |  |  |
|  |  | 11 | 4791 | 13 | 4703 | Normal | Normal |
|  |  | 9 | 8231 | 13 | 6574 |  |  |
|  |  | X | 3741 | Y | 2951 | Male | Male |
|  |  | X | 3320 | Y | 3589 |  |  |
|  |  | 3 | 4457 | X | 2141 |  |  |
|  |  | 18 | 6173 | X | 3423 |  |  |
| 116 | 39873 | 11 | 1613 | 21 | 2255 | Trisomy | Trisomy |
|  |  | 6 | 1345 | 21 | 1981 |  |  |
|  |  | 10 | 2164 | 18 | 1675 | Normal | Normal |
|  |  | 1 | 1246 | 18 | 1227 |  |  |
|  |  | 11 | 896 | 13 | 778 | Normal | Normal |
|  |  | 9 | 1947 | 13 | 1612 |  |  |
|  |  | X | 1561 | Y |  | Female | Female |
|  |  | X | 1045 | Y |  |  |  |
|  |  | 3 | 1048 | X | 982 |  |  |
|  |  | 18 | 1007 | X | 1024 |  |  |
| 117 | 39892 | 11 | 2247 | 21 | 2285 | Normal | Normal |
|  |  | 6 | 2424 | 21 | 2239 |  |  |
|  |  | 10 | 2685 | 18 | 2474 | Normal | Normal |
|  |  | 1 | 2565 | 18 | 2610 |  |  |
|  |  | 11 | 2235 | 13 | 2070 | Normal | Normal |
|  |  | 9 | 2299 | 13 | 2162 |  |  |
|  |  | X | 1307 | Y | 1213 | Male | Male |
|  |  | X | 1247 | Y | 1249 |  |  |
|  |  | 3 | 2140 | X | 1013 |  |  |
|  |  | 18 | 2324 | X | 1286 |  |  |
| 118 | 39943 | 11 | 3075 | 21 | 2964 | Normal | Normal |
|  |  | 6 | 2999 | 21 | 2976 |  |  |
|  |  | 10 | 3903 | 18 | 3100 | Normal | Normal |
|  |  | 1 | 2930 | 18 | 2843 |  |  |
|  |  | 11 | 2018 | 13 | 1942 | Normal | Normal |
|  |  | 9 | 3079 | 13 | 2614 |  |  |
|  |  | X | 3433 | Y |  | Female | Female |
|  |  | X | 2770 | Y |  |  |  |
|  |  | 3 | 2431 | X | 2266 |  |  |
|  |  | 18 | 2672 | X | 2757 |  |  |
| 119 | 39951 | 11 | 2135 | 21 | 2109 | Normal | Normal |
|  |  | 6 | 2048 | 21 | 1955 |  |  |
|  |  | 10 | 2454 | 18 | 2096 | Normal | Normal |
|  |  | 1 | 2537 | 18 | 2540 |  |  |
|  |  | 11 | 2044 | 13 | 1940 | Normal | Normal |
|  |  | 9 | 3345 | 13 | 2869 |  |  |
|  |  | X | 2165 | Y |  | Female | Female |
|  |  | X | 2391 | Y |  |  |  |
|  |  | 3 | 1544 | X | 1462 |  |  |
|  |  | 18 | 2846 | X | 2858 |  |  |
| 120 | 39954 | 11 | 2860 | 21 | 2804 | Normal | Normal |
|  |  | 6 | 2862 | 21 | 2988 |  |  |
|  |  | 10 | 3737 | 18 | 2954 | Normal | Normal |
|  |  | 1 | 2400 | 18 | 2464 |  |  |
|  |  | 11 | 1686 | 13 | 1582 | Normal | Normal |
|  |  | 9 | 2654 | 13 | 2144 |  |  |
|  |  | X | 1716 | Y | 1476 | Male | Male |
|  |  | X | 1087 | Y | 1068 |  |  |
|  |  | 3 | 2249 | X | 1062 |  |  |
|  |  | 18 | 2326 | X | 1262 |  |  |
| 121 | 40338 | 11 | 1423 | 21 | 1534 | Normal | Normal |
|  |  | 6 | 1559 | 21 | 1566 |  |  |
|  |  | 10 | 1815 | 18 | 1711 | Normal | Normal |
|  |  | 1 | 2246 | 18 | 2180 |  |  |
|  |  | 11 | 1721 | 13 | 1641 | Normal | Normal |
|  |  | 9 | 2502 | 13 | 2235 |  |  |
|  |  | X | 1878 | Y |  | Female | Female |
|  |  | X | 2061 | Y |  |  |  |
|  |  | 3 | 1609 | X | 1456 |  |  |
|  |  | 18 | 2323 | X | 2438 |  |  |
| 122 | 40393 | 11 | 2114 | 21 | 2140 | Normal | Normal |
|  |  | 6 | 2111 | 21 | 2001 |  |  |
|  |  | 10 | 2788 | 18 | 2301 | Normal | Normal |
|  |  | 1 | 1658 | 18 | 1693 |  |  |
|  |  | 11 | 1217 | 13 | 1193 | Normal | Normal |
|  |  | 9 | 2298 | 13 | 1886 |  |  |
|  |  | X | 2819 | Y |  | Female | Female |
|  |  | X | 1348 | Y |  |  |  |
|  |  | 3 | 1801 | X | 1605 |  |  |
|  |  | 18 | 1514 | X | 1548 |  |  |
| 123 | 40449 | 11 | 1000 | 21 | 1270 | Trisomy | Trisomy |
|  |  | 6 | 711 | 21 | 930 |  |  |
|  |  | 10 | 1369 | 18 | 1182 | Normal | Normal |
|  |  | 1 | 1159 | 18 | 1185 |  |  |
|  |  | 11 | 672 | 13 | 623 | Normal | Normal |
|  |  | 9 | 1533 | 13 | 1387 |  |  |
|  |  | X | 924 | Y |  | Female | Female |
|  |  | X | 895 | Y |  |  |  |
|  |  | 3 | 708 | X | 713 |  |  |
|  |  | 18 | 985 | X | 978 |  |  |
| 124 | 40077 | 11 | 4097 | 21 | 4309 | Normal | Normal |
|  |  | 6 | 5248 | 21 | 4901 |  |  |
| 125 | 40476 | 11 | 4549 | 21 | 4608 | Normal | Normal |
|  |  | 6 | 4855 | 21 | 4809 |  |  |
| 126 | 40486 | 11 | 2456 | 21 | 2458 | Normal | Normal |
|  |  | 6 | 1971 | 21 | 2145 |  |  |
|  |  | 10 | 2947 | 18 | 2563 | Normal | Normal |
|  |  | 1 | 2551 | 18 | 2747 |  |  |
|  |  | 11 | 1599 | 13 | 1602 | Normal | Normal |
|  |  | 9 | 2873 | 13 | 2420 |  |  |
|  |  | X | 1361 | Y | 1155 | Male | Male |
|  |  | X | 1099 | Y | 1065 |  |  |
|  |  | 3 | 1927 | X | 899 |  |  |
|  |  | 18 | 2237 | X | 1151 |  |  |
| 127 | 40498 | 11 | 398 | 21 | 375 | Normal | Normal |
|  |  | 6 | 363 | 21 | 352 |  |  |
|  |  | 10 | 512 | 18 | 465 | Normal | Normal |
|  |  | 1 | 522 | 18 | 469 |  |  |
|  |  | 11 | 292 | 13 | 278 | Normal | Normal |
|  |  | 9 | 567 | 13 | 485 |  |  |
|  |  | X | 534 | Y |  | Female | Female |
|  |  | X | 387 | Y |  |  |  |
|  |  | 3 | 286 | X | 240 |  |  |
|  |  | 18 | 351 | X | 348 |  |  |
| 128 | 40499 | 11 | 1266 | 21 | 1315 | Normal | Normal |
|  |  | 6 | 1266 | 21 | 1305 |  |  |
|  |  | 10 | 1605 | 18 | 1326 | Normal | Normal |
|  |  | 1 | 1613 | 18 | 1579 |  |  |
|  |  | 11 | 1122 | 13 | 1042 | Normal | Normal |
|  |  | 9 | 1673 | 13 | 1464 |  |  |
|  |  | X | 699 | Y | 616 | Male | Male |
|  |  | X | 675 | Y | 655 |  |  |
|  |  | 3 | 993 | X | 476 |  |  |
|  |  | 18 | 1597 | X | 793 |  |  |
| 129 | 40502 | 11 | 2509 | 21 | 2644 | Normal | Normal |
|  |  | 6 | 3171 | 21 | 3142 |  |  |
|  |  | 10 | 3261 | 18 | 2985 | Normal | Normal |
|  |  | 1 | 3311 | 18 | 3490 |  |  |
|  |  | 11 | 2440 | 13 | 2402 | Normal | Normal |
|  |  | 9 | 3355 | 13 | 3022 |  |  |
|  |  | X | 2764 | Y |  | Female | Female |
|  |  | X | 3063 | Y |  |  |  |
|  |  | 3 | 2712 | X | 2693 |  |  |
|  |  | 18 | 3297 | X | 3533 |  |  |
| 130 | 40512 | 11 | 5376 | 21 | 5453 | Normal | Normal |
|  |  | 6 | 4080 | 21 | 4002 |  |  |
|  |  | 10 | 5606 | 18 | 4692 | Normal | Normal |
|  |  | 1 | 3729 | 18 | 3723 |  |  |
|  |  | 11 | 2786 | 13 | 2624 | Normal | Normal |
|  |  | 9 | 3676 | 13 | 3294 |  |  |
|  |  | X | 2683 | Y | 2320 | Male | Male |
|  |  | X | 1860 | Y | 1915 |  |  |
|  |  | 3 | 3301 | X | 1580 |  |  |
|  |  | 18 | 3447 | X | 1902 |  |  |
| 131 | 40518 | 11 | 4393 | 21 | 4370 | Normal | Normal |
|  |  | 6 | 3223 | 21 | 3172 |  |  |
|  |  | 10 | 4340 | 18 | 3669 | Normal | Normal |
|  |  | 1 | 3876 | 18 | 3912 |  |  |
|  |  | 11 | 2248 | 13 | 2229 | Normal | Normal |
|  |  | 9 | 4513 | 13 | 3825 |  |  |
|  |  | X | 2190 | Y |  | Female | Female |
|  |  | X | 1909 | Y |  |  |  |
|  |  | 3 | 2018 | X | 2026 |  |  |
|  |  | 18 | 2099 | X | 2109 |  |  |
| 132 | 40534 | 11 | 3223 | 21 | 3090 | Normal | Normal |
|  |  | 6 | 2268 | 21 | 2293 |  |  |
|  |  | 10 | 3821 | 18 | 2935 | Normal | Normal |
|  |  | 1 | 3008 | 18 | 3212 |  |  |
|  |  | 11 | 2235 | 13 | 2026 | Normal | Normal |
|  |  | 9 | 2817 | 13 | 2534 |  |  |
|  |  | X | 3082 | Y |  | Female | Female |
|  |  | X | 3109 | Y |  |  |  |
|  |  | 3 | 2387 | X | 2346 |  |  |
|  |  | 18 | 2768 | X | 2707 |  |  |
| 133 | 40540 | 11 | 2684 | 21 | 2600 | Normal | Normal |
|  |  | 6 | 2239 | 21 | 2091 |  |  |
|  |  | 10 | 3275 | 18 | 2648 | Normal | Normal |
|  |  | 1 | 2862 | 18 | 2861 |  |  |
|  |  | 11 | 1898 | 13 | 1801 | Normal | Normal |
|  |  | 9 | 3374 | 13 | 2778 |  |  |
|  |  | X | 1320 | Y | 1075 | Male | Male |
|  |  | X | 1191 | Y | 1185 |  |  |
|  |  | 3 | 1861 | X | 971 |  |  |
|  |  | 18 | 2480 | X | 1280 |  |  |
| 134 | 40543 | 11 | 5141 | 21 | 5132 | Normal | Normal |
|  |  | 6 | 4512 | 21 | 4355 |  |  |
|  |  | 10 | 5370 | 18 | 4893 | Normal | Normal |
|  |  | 1 | 3909 | 18 | 4046 |  |  |
|  |  | 11 | 3022 | 13 | 2911 | Normal | Normal |
|  |  | 9 | 4190 | 13 | 3557 |  |  |
|  |  | X | 2941 | Y | 2566 | Male | Male |
|  |  | X | 2051 | Y | 2040 |  |  |
|  |  | 3 | 4340 | X | 2136 |  |  |
|  |  | 18 | 4188 | X | 2374 |  |  |
| 135 | 40934 | 11 | 1842 | 21 | 2118 | Normal | Normal |
|  |  | 6 | 2648 | 21 | 2301 |  |  |
|  |  | 10 | 2552 | 18 | 2369 | Normal | Normal |
|  |  | 1 | 2274 | 18 | 2278 |  |  |
|  |  | 11 | 2040 | 13 | 1876 | Normal | Normal |
|  |  | 9 | 2285 | 13 | 2015 |  |  |
|  |  | X | 1639 | Y | 1323 | Male | Male |
|  |  | X | 985 | Y | 980 |  |  |
|  |  | 3 | 2423 | X | 1202 |  |  |
|  |  | 18 | 2595 | X | 1421 |  |  |
| 136 | 40940 | 11 | 1347 | 21 | 1254 | Normal | Normal |
|  |  | 6 | 1230 | 21 | 1215 |  |  |
|  |  | 10 | 1567 | 18 | 1308 | Normal | Normal |
|  |  | 1 | 1589 | 18 | 1618 |  |  |
|  |  | 11 | 1197 | 13 | 1176 | Normal | Normal |
|  |  | 9 | 2325 | 13 | 2016 |  |  |
|  |  | X | 740 | Y | 608 | Male | Male |
|  |  | X | 616 | Y | 618 |  |  |
|  |  | 3 | 991 | X | 483 |  |  |
|  |  | 18 | 1494 | X | 733 |  |  |
| 137 | 40646 | 11 | 6707 | 21 | 6633 | Normal | Normal |
|  |  | 6 | 5148 | 21 | 5339 |  |  |
| 138 | 40886 | 11 | 6605 | 21 | 6578 | Normal | Normal |
|  |  | 6 | 4995 | 21 | 5067 |  |  |
| 139 | 40958 | 11 | 1446 | 21 | 1486 | Normal | Normal |
|  |  | 6 | 1411 | 21 | 1381 |  |  |
|  |  | 10 | 1829 | 18 | 1593 | Normal | Normal |
|  |  | 1 | 1273 | 18 | 1388 |  |  |
|  |  | 11 | 1017 | 13 | 919 | Normal | Normal |
|  |  | 9 | 1716 | 13 | 1379 |  |  |
|  |  | X | 940 | Y | 772 | Male | Male |
|  |  | X | 534 | Y | 546 |  |  |
|  |  | 3 | 1133 | X | 537 |  |  |
|  |  | 18 | 1398 | X | 733 |  |  |
| 140 | 45988 | 11 | 2679 | 21 | 2666 | Normal | Normal |
|  |  | 6 | 2154 | 21 | 2184 |  |  |
| 141 | 46537 | 11 | 3198 | 21 | 3317 | Normal | Normal |
|  |  | 6 | 2452 | 21 | 2358 |  |  |
| 142 | 46609 | 11 | 4797 | 21 | 5008 | Normal | Normal |
|  |  | 6 | 4489 | 21 | 4566 |  |  |

**Note:** The Serial No. and Sample ID columns provide unique identifiers for each sample. The Ref. Chr Name and its corresponding Peak Height represent the reference chromosome used for normalization in fragment analysis. The Target Chr Name and its Peak Height correspond to the chromosome analyzed for aneuploidy detection, with fluorescence intensity values obtained from the genetic analyzer. AI-Based Interpretation denotes the automated analysis performed using a Python-based artificial intelligence (AI) process, while Gold Standard (Karyotyping) indicates the chromosomal status determined through conventional karyotyping, which serves as the reference standard for validation.
